# The Epidemiology of *Plasmodium vivax* Among Adults in the Democratic Republic of the Congo: A Nationally-Representative, Cross-Sectional Survey

**DOI:** 10.1101/2020.02.17.20024190

**Authors:** Nicholas F. Brazeau, Cedar L. Mitchell, Andrew P. Morgan, Molly Deutsch-Feldman, Oliver John Watson, Kyaw L. Thwai, Andreea Waltmann, Michael Emch, Valerie Gartner, Ben Redelings, Gregory A. Wray, Melchior K. Mwandagalirwa, Antoinette K. Tshefu, Joris L. Likwela, Jess K. Edwards, Robert Verity, Jonathan B. Parr, Steven R. Meshnick, Jonathan J. Juliano

## Abstract

**Background:** Reports of *P. vivax* infections among Duffy-negative hosts have begun to accumulate throughout sub-Saharan Africa. Despite this growing body of evidence, no nationally representative epidemiological surveys of *P. vivax* in sub-Saharan Africa nor population genetic analyses to determine the source of these infections have been performed.

**Methods:** To overcome this critical gap in knowledge, we screened nearly 18,000 adults in the Democratic Republic of the Congo (DRC) for *P. vivax* using samples from the 2013-2014 Demographic Health Survey. Infections were identified by quantitative PCR and confirmed with nested-PCR. *P. vivax* mitochondrial genomes were constructed after short-read sequencing. Risk factors, spatial distributions and population genetic analyses were explored.

**Findings:** Overall, we found a 2.96% (95% CI: 2.28%, 3.65%) prevalence of *P. vivax* infections across the DRC. Nearly all infections were among Duffy-negative adults (486/489). Infections were not associated with typical malaria risk-factors and demonstrated small-scale heterogeneity in prevalence across space. Mitochondrial genomes suggested that DRC *P. vivax* is an older clade that shares its most recent common ancestor with South American isolates.

**Interpretation:** *P. vivax* is more prevalent across the DRC than previously believed despite widespread Duffy-negativity. Comparison to global and historical *P. vivax* sequences suggests that historic DRC *P. vivax* may have been transported to the New World on the wave of European expansion. Our findings suggest congolese *P. vivax* is an innocuous threat given its relatively flat distribution across space, lack of malaria risk factors, and potentially ancestral lineage.

**Funding:** National Institutes of Health and the Wellcome Trust.

## Introduction

*Plasmodium vivax* is the most prevalent malaria-causing parasite outside of Africa, accounting for approximately 14.3 million cases in 2017.^1^ The relative absence of *P. vivax* in Africa has long been attributed to the high prevalence of the Duffy-negative phenotype throughout most of sub-Saharan Africa (SSA).^2–4^ However, recent evidence has demonstrated that *P. vivax* infections are occurring throughout SSA among Duffy-negative hosts.^5^ Although these *P. vivax* infections have been associated with clinical cases, the distribution and extent of asymptomatic versus symptomatic disease in SSA remains unclear.^1,5,6^

Despite growing concern, no studies have systematically evaluated the burden, risk factors, spatial distribution, or origins of these SSA *P. vivax* infections. This lack of research is problematic as resources have begun to be directed towards diagnosing and addressing SSA *P. vivax*. While the return of *P. vivax* to SSA has the potential to undermine years of malaria control and elimination efforts, its threat-status has not yet been characterized. To address this critical gap in knowledge, we used samples from the Democratic Republic of the Congo (DRC) 2013-14 Demographic Health Survey (DHS) to screen a nationally representative population of nearly 18,000 adults for *P. vivax*. Surveys from the DHS program are community based and are expected to contain mostly healthy, asymptomatic participants. The DRC is situated in the center of Africa and is the second-largest country in SSA. Moreover, previous work has indicated that the DRC is a watershed region that links East and West Africa malaria.^7,8^ As a result, findings from the DRC may be generalizable to much of SSA.

Using this nationally representative survey, we provide the first national level estimate of *P. vivax* prevalence, associated risk factors, and the geographical distribution of cases in the SSA region. In addition, we use mitochondrial genomes to identify the potential origins of these infections. By coupling a nationally-representative, spatially-rich dataset with cutting edge spatial statistics, novel machine learning techniques, and genomics, we advance efforts to uncover the hidden distribution and history of *P. vivax* in SSA.

## METHODS

### Study Participants & Malaria Detection

We studied men and women aged 15 - 59 years and 15 - 49 years, respectively, that were surveyed in the 2013-2014 DRC DHS. Each participant answered an extensive questionnaire and provided a dried blood spot (DBS) for HIV and other biomarker screening. Spatial and ecological data were collected for each sampling cluster (Supplementary Text). We extracted DNA from each DBS using Chelex-100 (Bio-Rad, Hercules, CA) and Saponin and then screened all participants for *P. falciparum* using quantitative PCR (qPCR) targeting the *P. falciparum* lactate dehydrogenase gene as previously described.^9^ In addition, samples were screened for *P. vivax* using qPCR targeting the 18S ribosomal RNA gene.^10^ Samples that screened positive by 18S-qPCR underwent reflex confirmatory screening using a nested-PCR assay targeting 18S rRNA (Supplementary Text).^11^ To ensure the quality of DNA extraction, we excluded samples that failed to amplify human-beta-tubulin from analysis. Additionally, participants were excluded if they had missing data or were not a part of the DHS sampling schematic (Supplementary Figure 2).^12^ This study reanalyzes previously published *P. falciparum* data (sample size differences are due to different inclusion criteria).^9^

### Duffy Genotyping

Host Duffy antigen/chemokine receptor (DARC) genotype was determined using a previously validated High-Resolution Melt (HRM) assay.^13^ Genotypes that could not be definitely resolved by HRM were reconciled by Sanger sequencing.^6^ In addition, HRM results were validated by sequencing an approximately 10% random subset of samples (Supplementary Text).

### Risk Factor Modeling

*P. vivax* risk factors were identified from a comprehensive literature search and previous work from the 2013-2014 DRC DHS identifying *P. falciparum* risk factors.^9^ Risk factors were derived from the DHS questionnaires and other open-data sources (Supplementary Text). All continuous risk factors were standardized in order to promote model stability and ease of comparability. For dichotomized risk factors, the *a priori* protective level was selected as the referent level (e.g. HIV-negative) or the largest group if a protective level was not obvious (e.g. female for biological sex).

For each risk factor, confounding covariates were identified using a directed acyclic diagram built from our *a priori* causal framework of covariate and outcome relationships (Supplementary Figure 3). We then used inverse-probability weighting (IPW) to obtain marginal structural models and control for confounding between our risk factors and outcome of interest, malaria. IPWs were calculated with a super learning algorithm, which uses a loss-based approach with V-fold cross-validation to maximize predictions from an ensemble of candidate algorithms.^14^ We extended the standard super learning algorithm to account for spatial dependence among observations using spatial cross validation (Supplementary Text).^15^ The super learner algorithm was selected for IPW calculations to account for known issues and biases of functional form in fitting the exposure dose-response curve.^16^ Using the IPWs, we performed weighted regression using generalized estimating equations (GEE) with a logit-link function and binomial variance. IPWs and DHS sampling weights were accounted for in the GEE under the assumption that the distribution of sampling was independent of the distribution of confounding covariates, which allows for weights to be considered jointly: *w*_*f*_ = *w*_*s*_ * *w*_*iptw*_.

In addition, we considered several alternative explanations for the pattern of *P. vivax* infections. These alternative explanations included: (1) interactions between non-human ape (NHA) ranges and *P. vivax* cluster-level prevalences using permutation tests; (2) within-host interactions of *P. vivax* and *P. falciparum* using a multinomial likelihood-based model that assumes independent infection acquisition; and (3) *post-hoc* power calculations (Supplementary Text).

### *P. vivax* Prevalence Maps

We considered spatial autocorrelation with Moran’s I using a province adjacency matrix as well as a matrix of greater-circle distances between clusters.^17^ Greater circle distances were calculated using a geodesic approach.^18^ Significance was evaluated using a permutation test with 100,000 iterations and a one-sided p-value.

To determine the spatial distribution of *P. vivax*, we fit two types of Bayesian mixed spatial models: (1) a province-level areal model and (2) a cluster-level point process model. Province-based spatial models are important for intervention-planning, as most interventions in the DRC are implemented at the province-level. However, cluster-level models with Gaussian processes may be more representative of the intrinsic malaria distribution under the assumption of a continuous, and potentially heterogeneous, spatial process. Both sets of spatial models were fit with generalized linear mixed models using the logistic link function and a binomial error distribution with a spatial random effect (Supplementary Text). For each of the respective spatial-levels, we fit an intercept-only model and a model with all significant risk factors. Gelman’s deviance information criterion (DIC) was used to assess model fit.^19^

### *P. vivax* Mitochondrial Genomics

Three DRC *P. vivax* samples among children previously identified by our group from the 2013-2014 DRC DHS were considered to have the highest quality DNA and were prepared for sequencing.^20^ All analyses were limited to the mitochondrial genome (mtDNA) due to lack of coverage in the nuclear genome. Nucleotide variants were identified among all samples and used to create consensus haplotypes (Supplementary Text).

Using our three DRC isolates and 685 globally sourced sequences, we created subpopulations based on geographical K-means clustering. Genetic distance measures, phylogenetic trees, and genetic summary statistics were generated to explore population diversity and differentiation (Supplementary Text).

## RESULTS

### Study Population and Molecular Validation

Among the 17,972 samples successfully shipped to the University of North Carolina for processing, 17,934 (99·79%) were linked to the 2013-2014 DRC DHS survey. Of these 17,934 samples, 169 samples failed to amplify human beta-tubulin, 1,402 individuals had missing geospatial data, 237 individuals were classified as *de facto* (i.e. visitors rather than household members), 535 individuals had sampling weights of zero, and 17 individuals had missing covariate data and were excluded from analysis. In total, our final dataset consisted of 15,574 individuals across 489 clusters (Supplementary Figure 2).

Our *P. vivax* qPCR assay achieved an analytical sensitivity of 95% when at least 1·50×10^−7^ ng/μL of 18S target (approximately 7 parasites/μL) was present (Supplementary Table 2). No off-target amplification was observed when the qPCR assay was challenged with highly concentrated DNA template from other *Plasmodium* species (Supplementary Figure 1). *P. vivax* infections were confirmed by a separate, nested-PCR in 534 of 579 (93·6%) of qPCR-positive samples, with strong agreement between the initial and reflex confirmatory assays (Cohen’s = 0·80; Supplementary Text). All samples selected for Duffy-Genotyping validation had concordant HRM-qPCR and Sanger sequencing results, except for one sample that failed genotyping (Supplementary Text).

### Prevalence of *P. vivax* among Adults in the DRC

We restricted our prevalence estimates to 467 *P. vivax* infections that were confirmed by both qPCR and reflex nested-PCR (n_weighted_: 459·18, 95% CI_weighted_: 346·54, 571·82) and were among the 15,574 adults included in our study (n_weighted_: 15,490·20, 95% CI_weighted_: 14,060·60, 16,919·80). The national weighted prevalence of *P. vivax* among adults was 2·96% (95% CI_weighted_: 2·28, 3·65%), with cluster point-prevalences ranging from 0 - 46·15% (Figure 1). Most clusters only contained a single *P. vivax* infection, although the weighted count of infections ranged from 0 - 30·63 infections per cluster. In contrast, we identified 5,179 *P. falciparum* infections (n_weighted_: 4,651·94, 95% CI_weighted_: 4,121·93, 5,181·94) accounting for a weighted national prevalence of 30·03% (95% CI_weighted_: 27·87, 32·19%). Overall, there were 174 (n_weighted_: 145·29, 95% CI_weighted_: 108·11, 182·48) *P. falciparum* - *P. vivax* coinfections.

**Figure 1.**
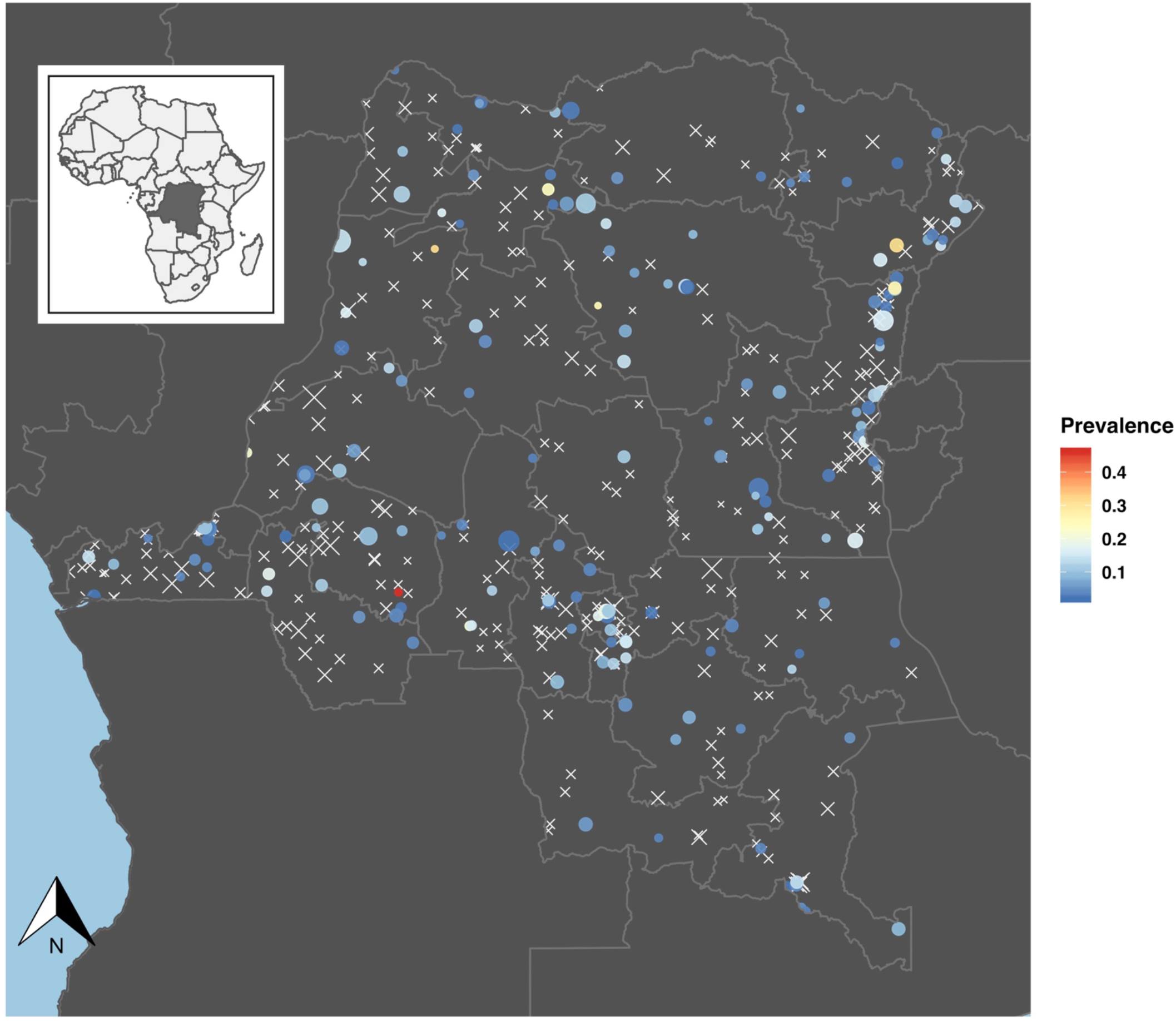
The Distribution of *P. vivax* Infections across the Democratic Republic of the Congo: For clusters with *P. vivax* infections, the prevalence is indicated by a blue-red spectrum, while the size of the point indicates the size of the cluster denominator. Clusters with no *P. vivax* infections are represented with white X-marks. *P. vivax* infections appeared to be diffusely spread throughout the country with cluster prevalences ranging from 0 - 46·15%.

### Risk Factors

Among the 579 qPCR-positive samples, only three had a putative Duffy-positive phenotype, all of whom were heterozygous at the loci associated with Duffy-negative phenotype (−33T:T/C). These three hosts (n_weighted_: 1·61, 95% CI_weighted_: −1·11, 4·34) were a part of the final study cohort, which led to an overall putative Duffy-positive phenotype frequency of 0·64% (Prevalence_weighted_: 0·35%, 95% CI_weighted_: 0·21, 0·58%) among those individuals infected with *P. vivax* included in our study. From our cross-species interference model that assumes independent acquisition of infections, we failed to find any significant interactions between *P. falciparum*-*P. vivax* co-infections (Supplementary Figure 5). Similarly, we did not find an association between NHA habitats and *P. vivax* cluster prevalence (Supplementary Figure 6). Although baseline characteristics differed by infection status, the differences appeared to be most pronounced between *P. falciparum* infections and uninfected-individuals rather than *P. vivax* infections and uninfected-individuals (Table 1).

**Table 1.**
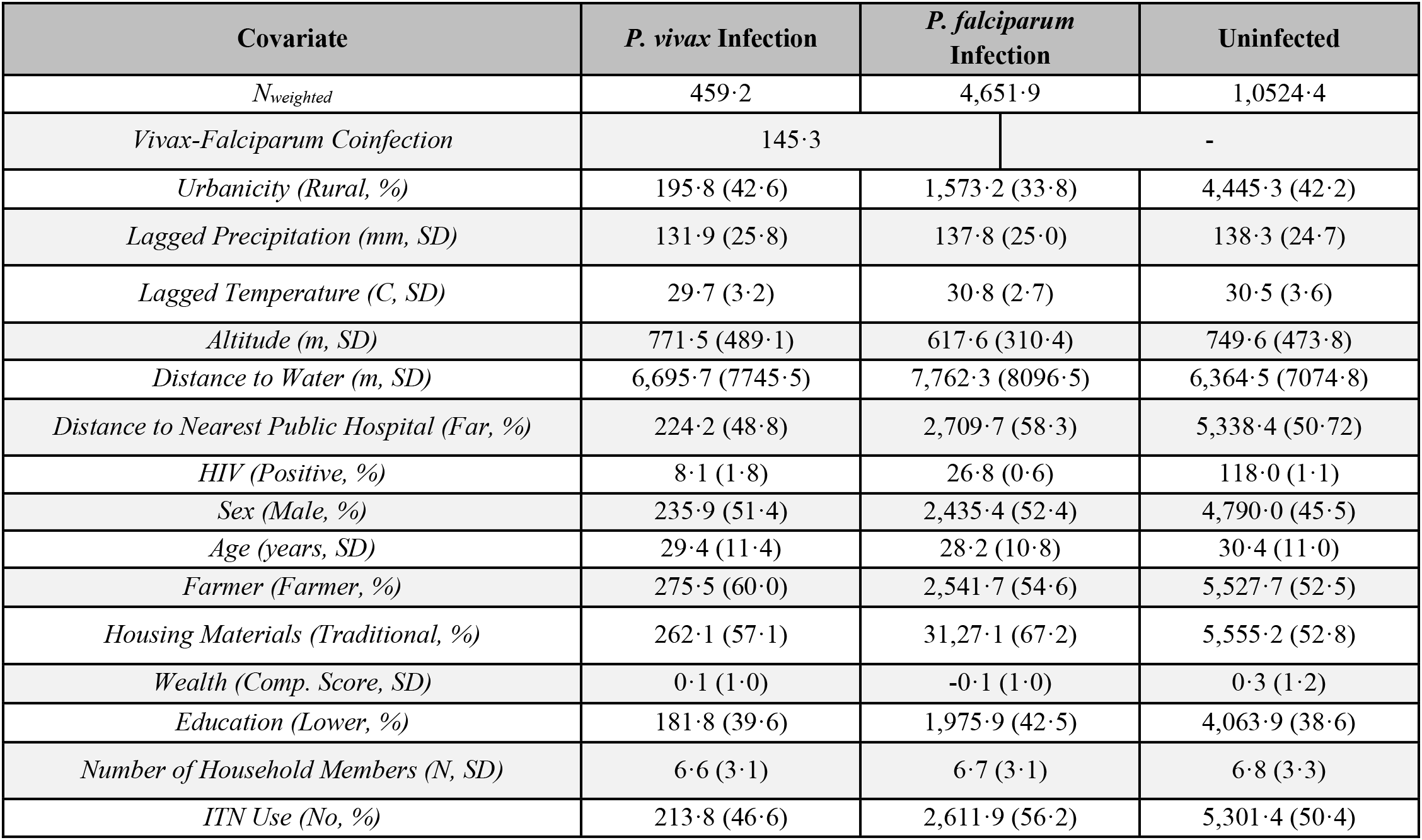
Baseline Distributions of Identified Risk Factors among Individuals with *P. vivax* Infections, *P. falciparum* Infections, and those that are Uninfected: Risk factor distributions appeared to differ by infection status, with more noticeable differences between *P. falciparum* infections and uninfected individuals. For dichotomized risk factors, the counts and percentages for each category are provided. For continuous risk factors, the mean and standard deviation (SD) are provided. *Abbreviations*: N - number of individuals, mm - millimeters, C – Celsius, m - meters, Comp. Score - composite score, ITN - insecticide-treated net.

In order to formally assess the risk-factor prevalence odds ratios (pORs) among *P. vivax* and *P. falciparum*, we adjusted for confounding using IPW. IPWs that were calculated with the spatially cross-validated super learner algorithm appeared to be stable with approximately log-normal distributions (Supplementary Figure 8). Additionally, for most covariates, IPW resulted in a considerable decrease in the average correlation among baseline covariates as compared to unadjusted baseline correlations (mean fold-reduction: 3·14, range: 0·85 - 7·63; Supplementary Figure 9).

When *P. vivax* was considered as the outcome of interest, higher-levels of precipitation were found to reduce prevalence (IPW-pOR: 0·79, 95% CI: 0·63, 0·99) while being a farmer appeared to increase prevalence (IPW-pOR: 1·42, 95% CI: 1·08, 1·89). In contrast, when considering *P. falciparum* infections as the outcome of interest, several risk factors were associated with prevalence: an urban setting reduced prevalence (IPW-pOR: 0·70, 95% CI: 0·54, 0·89), lack of insecticide-treated net (ITN) use increased prevalence (IPW-pOR: 1·23, 95% CI: 1·07, 1·42), increasing altitude reduced prevalence (IPW-pOR: 0·73, 95% CI: 0·65, 0·82), temperature increased prevalence (IPW-pOR: 1·41, 95% CI: 1·05, 1·90), lower levels of education increased prevalence (IPW-pOR: 1·44, 95% CI: 1·25, 1·67), higher levels of wealth reduced prevalence (IPW-pOR: 0·82, 95% CI: 0·73, 0·92), older age reduced prevalence (IPW-pOR: 0·81, 95% CI: 0·77, 0·86), while being male increased prevalence (IPW-pOR: 1·31, 95% CI: 1·20, 1·43).

Based on our *post hoc* power calculations for *P. vivax*, we were able to detect harmful pOR estimates of at least 1·54, 1·36, 1·30 with at least 80% power when the exposure probability was 10%, 25%, and 50%, respectively. In contrast, for *P. falciparum*, we were able to detect harmful pOR estimates of at least 1·18, 1·12, 1·10 with at least 80% power when the exposure probability was 10%, 25%, and 50%, respectively (Supplementary Figure 12).

### Spatial Distribution of *P. vivax*

When considering spatial autocorrelation, we found that the province-level showed a slight signal of structure for *P. vivax* prevalence (Moran’s I: 0·16; p = 0·05), but this structure did not hold at the cluster-level (Moran’s I: 0·02; p = 0.10). Among the *P. vivax* province-level models considered, we found that the best fitting model contained the precipitation, night light intensity, and farming covariates (Supplementary Table 7). Means-fitted province prevalences ranged from 1·10 - 7·42% (Figure 3A). Standard errors for the province prevalence estimates ranged from 4·44 × 10^−3^ - 2·76 × 10^−2^ (Supplementary Figure 11A).

**Figure 2.**
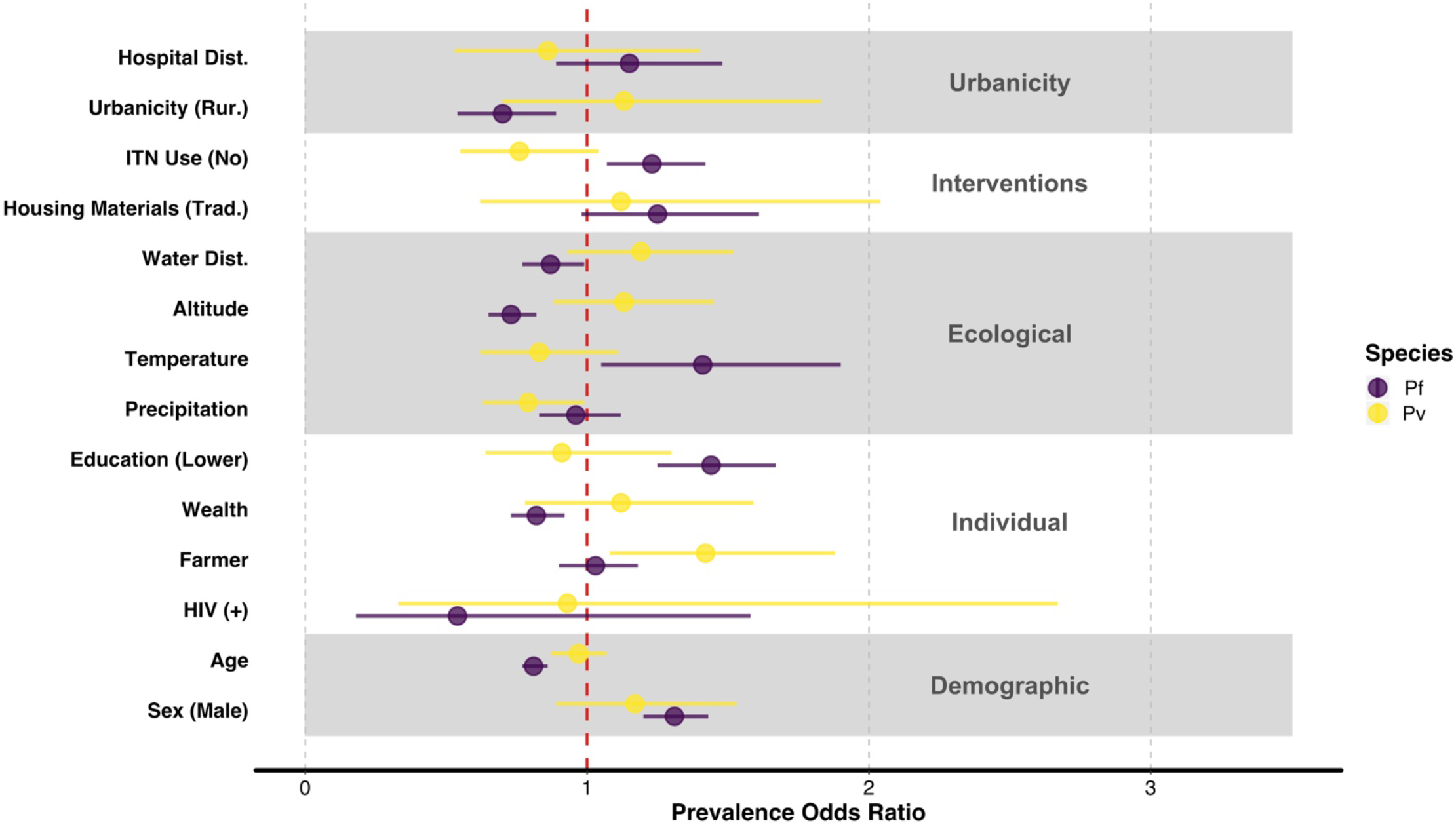
Inverse Probability Weight Adjusted Prevalence Odds Ratios for Expected Malaria Risk Factors: The inverse probability weight adjusted prevalence odds ratios (IPW-pORs) demonstrated a lack of risk factors for *P. vivax* infection, as all risk factors contain the null estimate (red line) with the exception of precipitation and farming. In contrast, numerous risk factors were associated with *P. falciparum*, including living in a rural area, ITN use, altitude, education, wealth, age, and biological sex. For both species, hospital distance, traditional housing materials, distance to water, and HIV-status were not significant risk factors. The unadjusted pORs effect estimates and confidence intervals as well as the IPW-pORs are provided in Supplementary Table 6 for reference. *Abbreviations*: Hospital Dist. – Distance to a hospital, Water Dist. – Distance to water, Rur. - rural, Trad. - traditional, ITN - insecticide-treated net.

**Figure 3.**
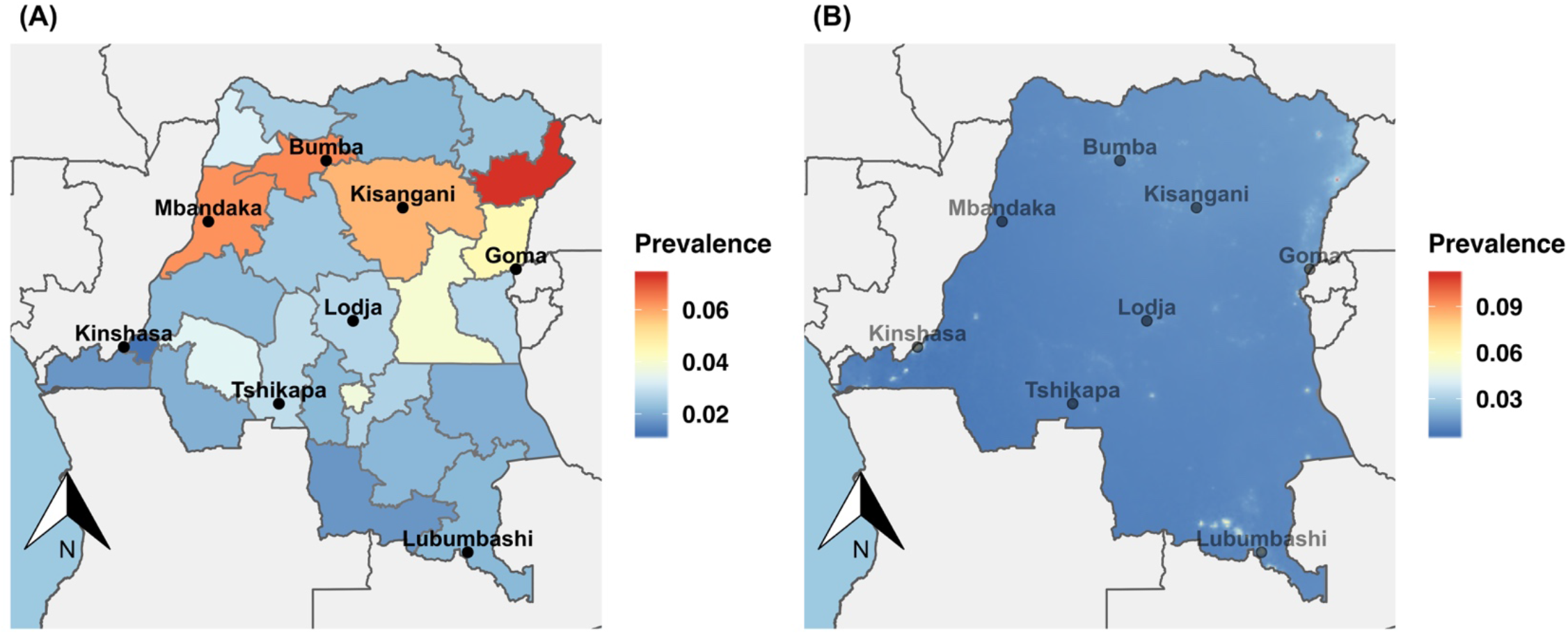
Spatial Model Posterior Means: The means of the posterior prevalence distribution for the province-level (left) and cluster-level (right) models. At the province-level, *P. vivax* infections appeared to be more common in the north. However, this process was not recapitulated at the cluster-level, where very local transmission appeared to dominate with a few focal regions of high prevalence amidst a relatively uniform background of *P. vivax* prevalence.

Similarly, when we modeled the spatial distribution of *P. vivax* at the cluster-level, we found that the best fitting model contained the precipitation, night light intensity, and farming covariates (Supplementary Table 7). Based on the model predictions, *P. vivax* fitted prevalence ranged from 0·50 - 11·20% across the DRC (Figure 3B). The standard errors around the prevalence predictions ranged from 1·62 × 10^−8^ - 2·30 × 10^−6^ (Supplementary Figure 11B). Most *P. vivax* prevalence predictions were less than the observed national prevalence (19,903/20,000; 99·52%).

### *P. vivax* Diversity, Differentiation, and Phylogeography

We achieved high-quality coverage in ≥98·0% of the mtDNA, with an average mtDNA base-depth of 40·85 among the three sequenced DRC *P. vivax* samples. Among the 636 publicly available Illumina sequenced *P. vivax* isolates that passed QC-thresholds, we detected 57 unique mitochondrial haplotypes (Supplementary Figure 13). Among the haplotypes, we identified 65 biallelic sites and 1 polyallelic site, with most mutations occurring in the non-protein coding regions (Ti:Tv = 1·2). Overall, the NHA and the Asian *P. vivax* population demonstrated the greatest within-population nucleotide and haplotype diversity, while there was limited within-population diversity among the isolates from the DRC (Supplementary Table 9). Based on between-population measures of nucleotide diversity, the DRC samples were most similar to samples from the Americas. However, when considering pairwise measures of F_st_ between populations, the DRC population appeared to be relatively isolated from other populations (Supplementary Table 10). When considering the evolutionary relationship of the DRC samples with samples sourced from across the globe, we found that the DRC samples formed a separate monophyletic clade (Bootstrap Support: 62·0%). The DRC monophyletic clade had a most recent common ancestor (MRCA) with a clade that contained a subset of samples from Peru (Figure 4; Bootstrap Support 11·8%). Although the DRC haplotype appeared to be most closely related to haplotypes circulating in the Americas, the DRC haplotype was similar to haplotypes from the Asian and Oceanic populations (Figure 5).

**Figure 4.**
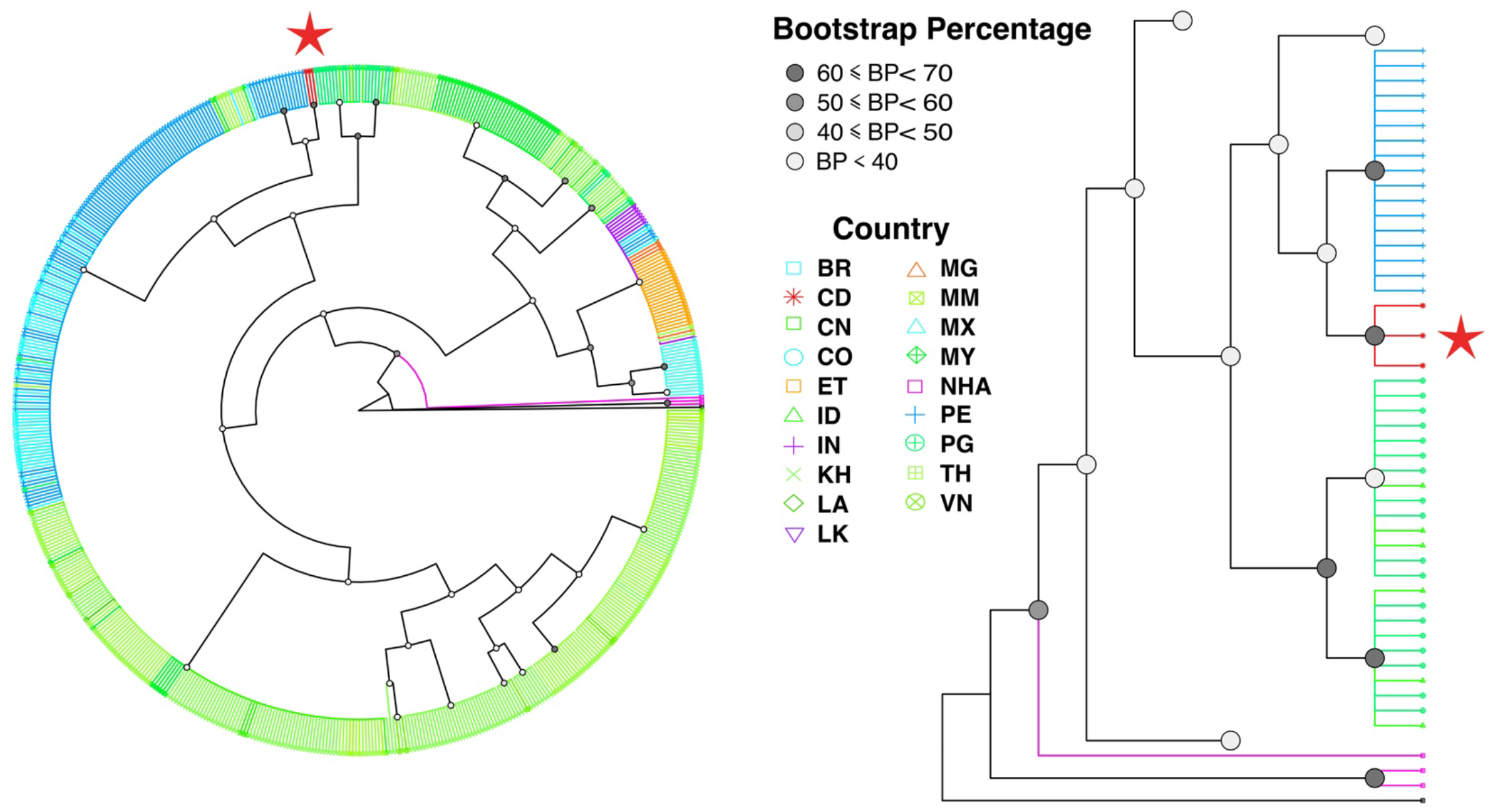
Phylogenetic Tree of *P. vivax* Global Isolates: Comparison of DRC *P. vivax* with 636 globally sourced *P. vivax* isolates showed that *P. vivax* shared a most recent common ancestor with samples from Peru. Although the DRC-Peru node did not have strong support, collapsing the node still results in the same conclusion of *P. vivax* sharing a most recent common ancestor with isolates from South America. The full phylogenetic tree is provided (left), with various clades not along the DRC ancestry collapsed for a focused view of the DRC clade (right). Isolates from the Americas are colored in shades of blue and include Brazil (BR), Colombia (CO), Mexico (MX), and Peru (PE). Asian countries are indicated in shades of green and include China (CN), Indonesia (ID), Cambodia (KH), Laos (LA), Myanmar (MM), Malaysia, Papua New Guinea, Thailand (TH), and Vietnam (VN). India (IN) and Sri Lanka (LK) are indicated in shades of purple, while Ethiopia (ET) and Madagascar (MG) are indicated in shades of orange. Finally, the Democratic Republic of the Congo (CD) is shown in red and non-human apes (NHA) are shown in magenta. *P. cynomologi* was set as the tree root and is indicated in black.

**Figure 5.**
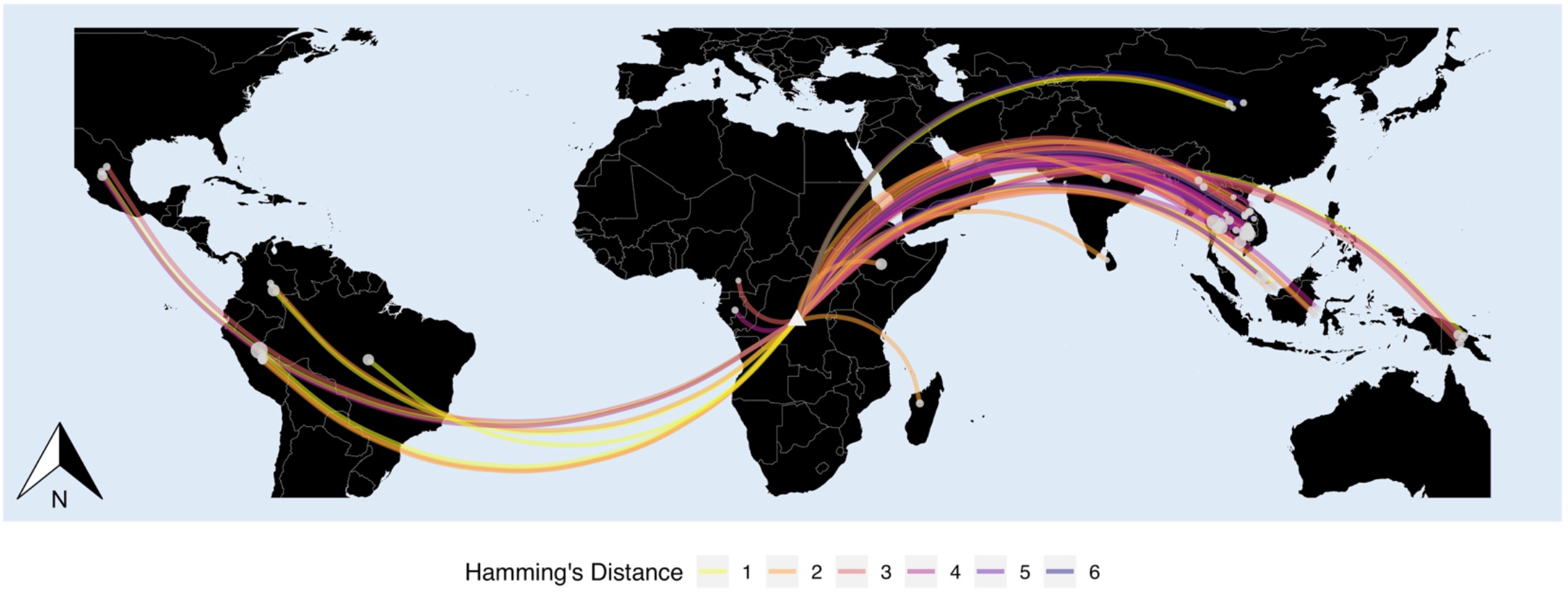
Haplotype Genetic Distances among Global Isolates with respect to the DRC: Although DRC samples may share a most recent common ancestor with isolates from South America, DRC haplotypes contain elements from both South America, Asia, and the Oceanic regions. Given that DRC haplotypes are more similar to haplotypes in South America and Asia than in Africa, this suggests that the DRC *P. vivax* may be an ancestral strain that potentially seeded those regions. All unique haplotypes with respect to country of origin are provided for comparison and context (Supplementary Figure 13).

## DISCUSSION

*P. vivax* infections among adults in the DRC are more common than previously realized. From our spatially robust dataset across the DRC, we detected 467 *P. vivax* infections corresponding to a national prevalence of 2·96% (95% CI_weighted_: 2·28, 3·65%). Among those infected, nearly all were Duffy-negative (576/579, 99·48%).

Malaria risk-factors typically associated with *P. falciparum* infection, such as ITN use and wealth, were not associated with *P. vivax* infection. Instead, only precipitation and farming were identified as *P. vivax* risk factors. This relationship between *P. vivax* prevalence and precipitation has been previously described, although the underlying effect is likely complicated by other ecological factors, such as vector habitats, seasonality, altitude, and temperature.^21^ Similarly, increased prevalence of malaria has previously been attributed to agriculture and farming in the DRC.^22^ Overall, the *P. vivax* malaria risk factors differed greatly from risk factors found for *P. falciparum* in this study using the same methodological approach. This contrast between *P. vivax* and *P. falciparum* risk factors may be the result of the shortened intrinsic period in *P. vivax* or hypnozoite infections being resilient to typical antimalarial interventions.^23^

*P. vivax* infections were found throughout the entire country with a few focal regions of relatively high prevalence. The highest prevalence of *P. vivax* was found in the Ituri province. This may be due to cross-border migration with South Sudan and Uganda, which border countries that are endemic for *P. vivax* (*P. vivax* infections have been reported in both countries).^1,5^ However, the sources of disease in other provinces are not clear. Microheterogeneity in *P. vivax* prevalence has previously been reported in the Amazon and was found to be associated with human movement ^24^. Future *P. vivax* epidemiological studies in the DRC should consider human mobility data, particularly with respect to Kinshasa and regions along the eastern border where interactions with Duffy-positive individuals may be more frequent.

Although there appears to be small-scale heterogeneity of *P. vivax* in the DRC, more than half of predicted prevalences were less than one-percent and 99·95% of predicated prevalences were less than the observed national average. These localized regions of prevalence, or “hotspots,” contrast the broad spatial distribution of *P. falciparum* infections previously observed in the 2007 and 2013 DRC DHS.^9,25^ As a result, we suggest that *P. vivax* has been unable to gain a foothold in the region and is persisting rather than spreading.

The relatively large differences in our DRC *P. vivax* and the NHA mitochondrial genomes likely negates recent zoonotic transmission as the source of DRC *P. vivax*. We identified a MRCA between the DRC samples and a subset of samples from Peru during phylogenetic analysis. Although the node support for the DRC-Peru relationship was weak (11·6%), collapsing the node does not alter the conclusion that the DRC MRCA was most similar to extant South American parasites. This finding that Africa may have seeded American *P. vivax* has recently gained traction based on analysis of a historical sample originating from the Ebro Delta in Spain, circa 1944.^26–28^ Using this historical sample, the authors demonstrated that now extinct European *P. vivax* was closely related to extant *P. vivax* from the Americas, potentially dating to the European colonial expansion during the 15th century.^26,27^ When we included the Erbo-1944 sample in our comparisons, we found that the mitochondrial haplotype differed by only a single base-pair from our DRC haplotypes. As a result, we hypothesize that DRC *P. vivax* may have migrated from Africa to Europe prior to being transported to the New World on the wave of European expansion.

However, the history of DRC *P. vivax* is not straightforward. The haplotypes differed from samples collected in Asia, Oceania, and the Americas by only a single base pair. This close relationship may indicate ongoing or historical mixing with Asian and Oceanic *P. vivax*, the latter idea supported by the genetic measures of population differentiation. In addition, the Erbo-1944 consensus haplotype also matched haplotypes from the Americas, Asia, and Oceania populations. These similarities may have arisen due to waves of historical introgression and panmixia among *P. vivax* globally or may be an artifact of our conservative approach to variant filtering. Consistent with previous reports, we found relatively few informative sites in the *P. vivax* mitochondria.^28^ Despite this low variation, the mitochondria is a non-recombining region with putatively neutral SNPs that is ideal for phylogenetic analysis to resolve ancestry.^28^

The DRC is a critical region for the study of malaria in SSA due to its size, central location, and the indication that it bridges East and West Africa malaria.^7^ These characteristics allow the DRC to serve as a microcosm of the region.^7,8^ The main limitations of our study are the cross-sectional design, which limits inference of effects with a temporal component (e.g. seasonality) and restricts the study population largely to asymptomatic individuals, and the small number of high-quality DRC mitochondrial sequences generated. Future efforts will require more biological material than DBS to improve the likelihood of successful genomic sequencing. With whole genomes, signatures of selection for adaptation to the Duffy-negative host could be evaluated alongside robust demographic inference.

Until recently, *P. vivax*, was an unrecognized cause of disease in SSA. This study provides the first systematic and nationally representative survey of *P. vivax* in a SSA country not considered endemic for the disease. We demonstrated that *P. vivax* is circulating at prevalences higher than previously thought, despite a high frequency of Duffy-negativity.^1^ However, *P. vivax* infections were not associated with classic malaria risk factors, were spread diffusely throughout the country, and may represent an old lineage. These findings suggest that *P. vivax* may have been circulating in SSA as an innocuous, chronic infection that was overlooked in past studies due to frequently sub-microscopic or low parasitemia infections. This hypothesis is consistent with previous work that suggests *P. vivax* infections among Duffy-negative individuals are frequently mild and asymptomatic compared with Duffy-positive individuals.^5,6^ Finally, emerging research suggests that genotypically Duffy-negative hosts express the Duffy antigen among erythroid progenitors in the bone marrow and that *P. vivax* gametocytes are able to mature and proliferate in the bone marrow of non-human primate animal models.^29,30^ Collectively, this suggests that *P. vivax* in sub-Saharan Africa may be persisting as low parasitemic, asymptomatic, or relatively innocuous infections, by hiding in the bone marrow of Duffy-negative hosts. While the malaria community should remain mindful of *P. vivax* in SSA, its distribution and low prevalence support continued investments targeting *P. falciparum* as likely having the greatest impact on malaria elimination, morbidity, and mortality.

## Data Availability

Pending

## ACKNOWLEDGEMENTS

The authors would like to thank the congolese survey field teams and study participants. The authors also thank the numerous open source data platforms that were used in this study, including the Demographic Health Survey from the Democratic Republic of the Congo 2013-2014, Open Street Map (map data copyrighted OpenStreetMap contributors and available from https://www.openstreetmap.org), Database of Global Administrative Areas, Level-1 and Atmosphere Archive & Distribution System Distributed Active Archive Center, the Earth Observations Group at National Oceanic and Atmospheric Administration/National Centers for Environmental Information, European Space Agency, and Copernicus Atmosphere Monitoring Services. We also thank the European Nucleotide Archive and previous authors for making their next generation sequencing data available to the public. Finally, the authors would like to thank Sandra Mendoza Guerrero for help with the hybrid captures.

## AUTHOR CONTRIBUTIONS

NFB designed experiments, conducted laboratory work, conducted analysis and wrote the manuscript. CLM conducted laboratory work, advised on analyses, and participated in manuscript preparation. APM, OJW, MDF, VG, BR, JE, RV, helped develop software, advised on analyses, and participated in manuscript preparation. MKM, AKT, JLL, collected samples and participated in manuscript preparation. AW and KLT conducted laboratory work and participated in manuscript preparation. ME, GW, JBP, SRM, and JJJ helped conceive the study, contributed to the experimental design, advised on analyses, and participated in manuscript preparation.

## ETHICAL APPROVALS

This study was approved by the University of North Carolina at Chapel Hill IRB.

